# Non-pharmacological measures for the treatment of iron deficiency anemia: A systematic review and meta-analysis

**DOI:** 10.1101/2022.04.23.22274204

**Authors:** Francisca Mayara Brasileiro Gomes, Francisco Plácido Nogueira Arcanjo, Francisco Leandro Fonteles Moreira, Ivana Cristina de Holanda Cunha Barreto, Luiz Odorico Monteiro de Andrade, Thiago Corrêa de Oliveira, Eliana Pereira Vellozo, Benjamin Israel Kopelman, Cecília Costa Arcanjo Freire, Caio Plácido Costa Arcanjo, Maria Aparecida Zanetti Passos

## Abstract

**Objective:** To conduct a systematic review and meta-analysis to evaluate the effectiveness of non-pharmacological measures for the treatment of iron-deficiency anemia (IDA).

**Data sources:** MEDLINE (via PubMed), Cochrane library, SciELO/LILACS and EMBASE up to June 2021

**Study selection and data extraction:** We identified all randomized controlled trials (RCTs) that used non-pharmacological measures to treat IDA including iron pots/ingots, or food use were included. The outcomes of interest were hemoglobin (Hb) concentrations and prevalence of anemia.

**Results:** 479 studies were retrieved from the databases, of which 4 duplicate records were removed. After, all titles and abstracts were reviewed, 23 articles were considered potentially relevant, and were read in full and checked for eligibility. Three articles met all inclusion criteria. We also conducted a manual search for citations and a further 8 records were identified and checked for eligibility. Eleven RCTs were included in this review. Estimates showed that the use of non-pharmacological measures was associated with a statistically significant overall increase in mean Hb (MD +0.45 g/dL, 95% CI 0.05 to 0.85, p=0.03). The effect of non-pharmacological measures on the prevalence of IDA was analyzed in only 5 RCTs. Participants in the intervention groups were 2.78 times less likely to suffer from IDA than those in the control groups, OR=2.78, 95% CI 0.93, 8.29, however without significance for the overall effect (p=0.07).

**Conclusion:** Non-pharmacological therapies have a positive effect on iron balance, and can a useful adjunct to programs to prevent and treat IDA in at-risk populations. (PROSPERO registration number CRD42021261773).

## Introduction

Anemia is an important indicator of malnutrition and health with great consequences for socioeconomic development. Children under two years of age with severe anemia are at higher risk of mortality, and even mild forms, which can be corrected, cause permanent cognitive damage by decreasing attention span and memory deficits.^1,2^.

According to WHO estimates, between 1993 and 2005, anemia affected approximately a quarter of the world’s population, which then corresponded to 1.62 billion people, the majority being children under the age of four years. A recent study showed a decrease in the prevalence of anemia between 1990 and 2010 from 40.2% to 32.9% of the world population, especially among men. The regions most affected by anemia were Southeast Asia and sub-Saharan Africa. Iron deficiency (ID) was the main cause, and children under the age of 5 years were most affected. Malaria, schistosomiasis and chronic renal failure were the causes of anemia for which prevalence most increased in this period.^3^

The Pan American Health Organization states that Brazil is in second place among countries in Latin America and the Caribbean with the highest prevalence of anemia (30%); only Peru has higher numbers with 57%.^4,5^ According to the Brazilian Ministry of Health, in 2006, the general estimate of the prevalence of anemia in the country was 25 to 30%. In Brazil there is no official record of the prevalence of the disease; only regional studies can be found.^6-9^

Among the causes of anemia, ID is the most common, accounting for 50% of cases. In developing countries such as Brazil, diet has an important role in anemia caused by ID, mainly because iron, although present in cereals and legumes (foods that are easily accessed), its presence is in a low availability form. Therefore, in risk groups, it is necessary to stimulate the consumption of animal-based foods like meat and chicken, which contain an optimal amount of iron in its active form, to prevent iron-deficiency anemia (IDA).^5,8,10-13^

Currently, ID in Brazil is more common than other deficiencies, such as hypovitaminosis A, primary iodine deficiency, or protein-energy malnutrition. However, ID may also be the result of blood loss. While menstrual loss is the major non-nutritional cause of ID in women of childbearing age, in men and postmenopausal women, bleeding from the gastrointestinal tract is frequent cause.^11,14^

Even today, despite numerous health policies developed by the WHO and other agencies, anemia is a highly prevalent pathology in Brazil and worldwide. It is important not to undervalue or trivialize the disease, even when it is mild and oligosymptomatic, given that socioeconomic, intellectual and quality of life losses may result from this problem. This is without considering the increase in mortality in biologically more fragile populations.^15^

A nutrition education strategy that aims at the adequate quantitative and qualitative consumption of foods that are sources of different nutrients is an alternative that has low cost and does not produce undesirable effects. Through this, it is possible to increase the population’s knowledge about ID and clarify about monotonous and iron-poor diets, which constitute one of the main causes of this deficiency. It is noteworthy, however, that changes in eating habits are not quickly achieved, making the strategy effective in the long term. Intervention studies through educational actions aimed at parents of children under the age of 24 months were effective in preventing ID. These results confirm that adherence to correct dietary practices is important to address this problem, which presents high prevalence in this age group.^16^ However, for positive results, their actions must guarantee the consumption of foods rich in iron and dietary strategies that increase the bioavailability of iron in the diet, in addition to reducing the factors that hinder it.

It is still uncertain whether non-pharmacological measures, including iron-rich foods, are able to prevent and treat anemia in vulnerable populations. Therefore, this study intends to evaluate the effectiveness of non-pharmacological measures for the prevention and treatment of IDA.

## Methods

### Study design

This is a systematic review and meta-analysis to verify the efficacy and safety of non-pharmacological measures for the treatment and prevention of IDA.

### Definition of the clinical question

The definition of the specific question was carried out under the acronym PICOS, as shown in Figure 1.

**Figure 1.**
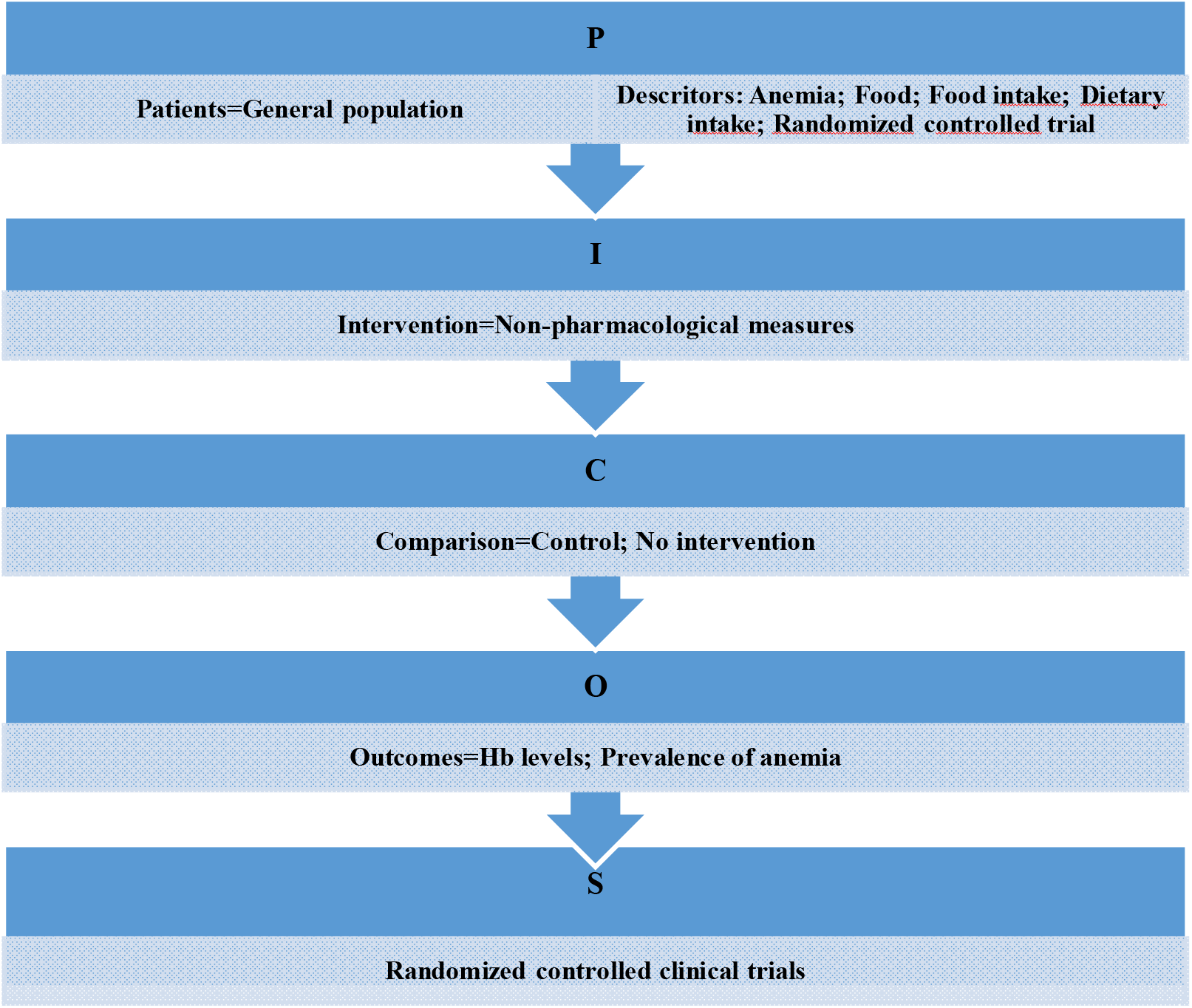
PICOS Question.

### Information sources

To conduct this study, the MEDLINE (via PubMed), Cochrane library, SciELO/LILACS and EMBASE databases were used. They were searched with no date or language restrictions. In addition, the reference list of included studies was also manually searched by the reviewers. This review was designed and conducted in accordance with the PRISMA^17^ guidelines and registered at PROSPERO under the registration number CRD42021261773.

### Search

To retrieve the articles (last search date was June 2021) we used the Boolean operators OR and AND. For the SciELO/LILACS database, the operators (Anemia) AND (Food) were used. In the Cochrane library the search algorithms were (Anemia), (Food Intake), (Randomized Controlled Trial) inserted without modification. However, to perform the search in the MEDLINE (via PubMed) and EMBASE databases, the terms MeSH and EMTREE were inserted, respectively.

Therefore, to perform the PubMed search, the appropriate MeSH terms were searched first. When inserting the studied condition (Anemia) the platform generated the following term: “Anemia”. For the term (Food Intake), the following were generated: (Intake, Food) OR (Nutrient Intake) OR (Intake, Nutrient) OR (Nutrient Intakes) OR (Nutritional Intake) OR (Intake, Nutritional) OR (Nutritional Intakes) OR (Dietary Intake) OR (Dietary Intakes) OR (Intake, Dietary) OR (Micronutrient Intake) OR (Intake, Micronutrient) OR (Micronutrient Intakes) OR (Ingestion) OR (Feed Intake) OR (Feed Intakes) OR (Intake, Feed) OR (Macronutrient Intake) OR (Intake, Macronutrient) OR (Macronutrient Intakes) OR (Calorie Intake) OR (Calorie Intakes) OR (Intake, Calorie) OR “Eating”[MeSH].

A randomized controlled trial filter^18^ was used with the previously described MeSH terms: (randomized controlled trial [pt] OR controlled clinical trial [pt] OR randomized controlled trials [mh] OR random allocation [mh] OR double-blind method [mh] OR single-blind method [mh] OR clinical trial [pt] OR clinical trials [mh] OR (“clinical trial” [tw]) OR ((singl* [tw] OR doubl* [tw] OR trebl* [tw] OR tripl* [tw]) AND (mask* [tw] OR blind* [tw])) OR (“latin square” [tw]) OR placebos [mh] OR placebo* [tw] OR random* [tw] OR research design [mh:noexp] OR follow-up studies [mh] OR prospective studies [mh] OR cross-over studies [mh] OR control*[tw] OR prospectiv* [tw] OR volunteer* [tw]) NOT (animal [mh] NOT human [mh]).

To search the EMBASE platform, in addition to the terms listed above, the EMTREE terms were used: ‘iron deficiency anemia’/exp AND ‘iron deficiency’/exp AND (‘food intake’/exp OR ‘appetite regulation’ OR ‘feed intake’ OR ‘feeding methods’ OR ‘food consumption’ OR ‘food intake’ OR ‘food intake’ OR ‘food intake regulation’ OR ‘food uptake’ OR ‘meal’.

In addition to this search strategy, a manual search was carried out to analyze the references cited in the selected articles to identify additional documents for review.

### Study selection

Randomized controlled clinical trials (RCTs) that used non-pharmacological measures to treat IDA including iron pots, iron ingots, or food use were included. There was no age group restriction. We excluded studies that did not include any quantifiable data.

### Data collection process and quality assessment

Data were extracted from the included studies in standardized tables by one reviewer and verified by a second. Data were collected on study title, 1^st^ author, year of publication, country, objectives, number of participants, population, type of intervention and comparison, primary and secondary outcomes, and main results. Synthesized data from all studies included in this review are shown in Table 1.

**Table 1.**
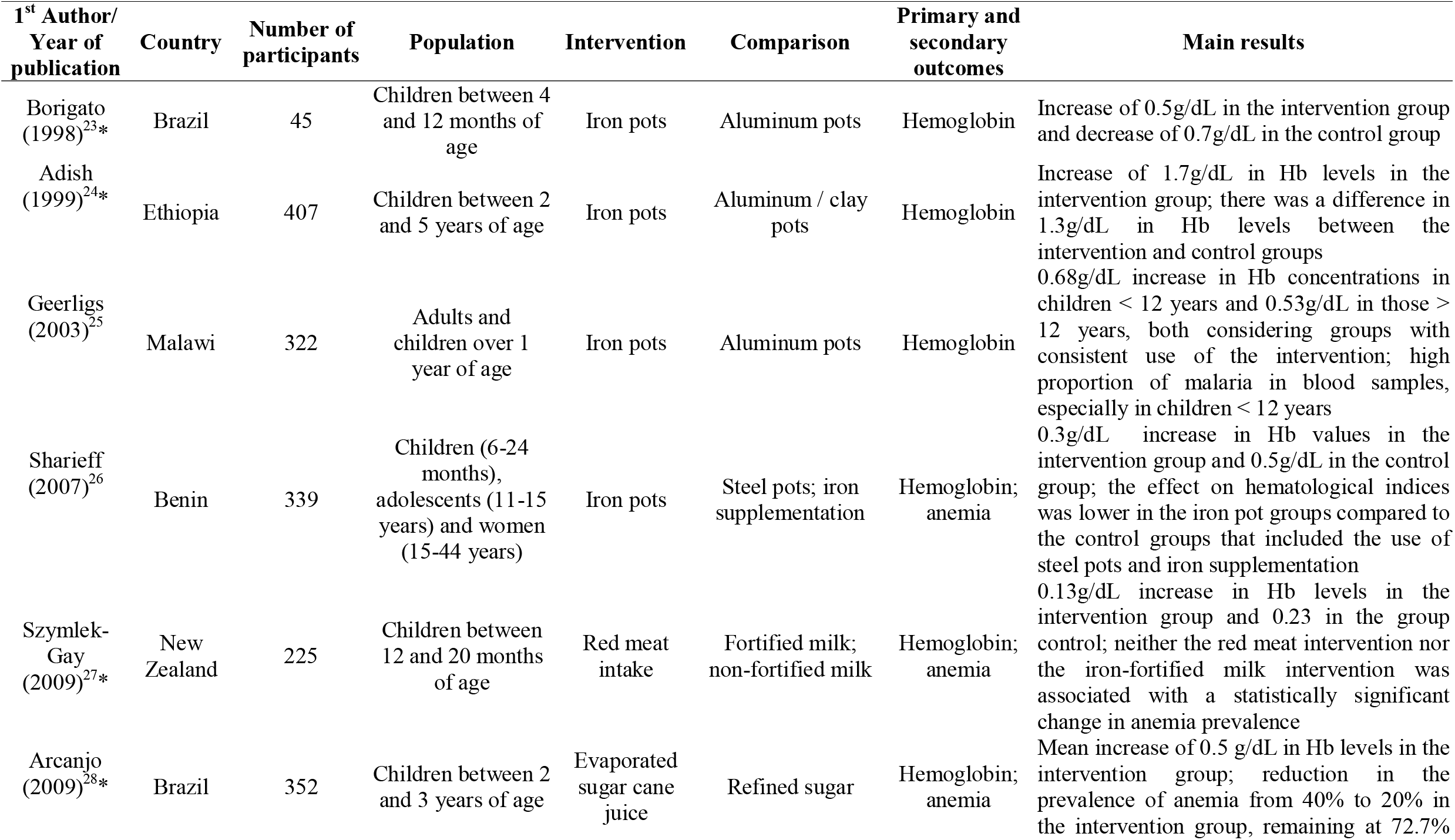
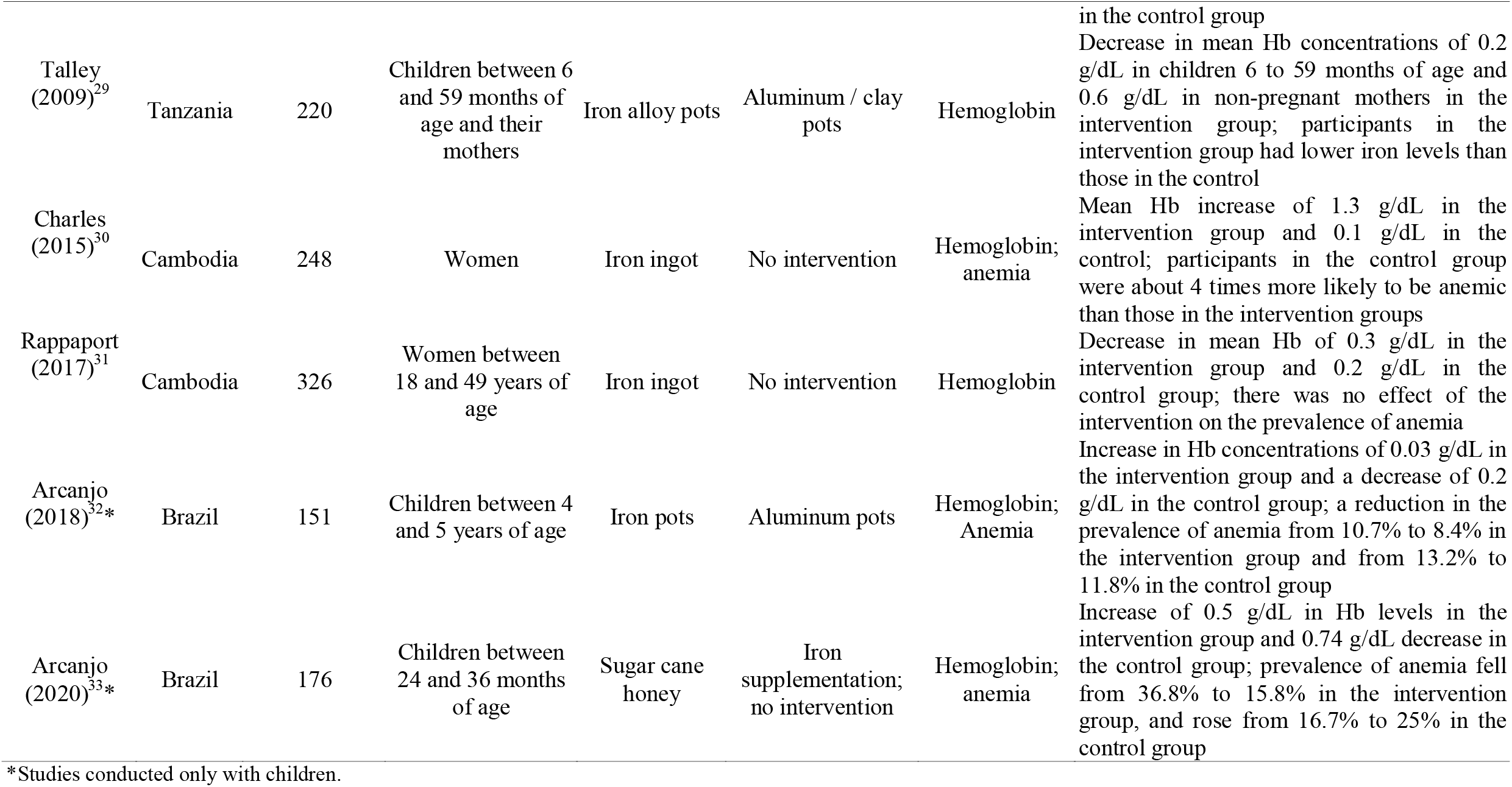
Characterization of included randomized clinical trials.

The Cochrane Risk-of-Bias (RoB 2.0) tool (available via the Risk of Bias tools website: www.riskofbias.info) was used to assess the quality and risk of bias in the RCTs included in this review. This tool is structured into five domains, through which bias may be introduced into the result: 1) bias arising from the randomization process - used to generate the participants’ allocation sequence, which should be random; 2) bias due to deviations from intended interventions – this domain concerns the patient and the study team not knowing (being “blind”) to which group the patient was allocated and whether there were deviations from the proposed intervention that could affect the outcome; 3) bias due to missing outcome data - loss to follow-up of study participants and, in case of loss, the reason for its occurrence; 4) bias in measurement of the outcome – outcome variable assessment, participant, researcher or data collector do not know which group the participants have been assigned to; and 5) bias in selection of the reported result - the possibility that the researchers assessed outcomes through multiple assessments, but reported only the most convenient one(s).

To assess the quality of evidence, the Grading of Recommendations Assessment, Development and Evaluation (GRADE)^20^ was used, thus ensuring a systematic and transparent process in this systematic review process. This system classifies evidence into high, moderate, low or very low quality, considering all the factors that determine how reliable the results are. Two outcomes were evaluated, hemoglobin (Hb) concentrations, and prevalence of anemia.

### Synthesis of results

Data was summarized and aggregated in a quantitative synthesis of the studies through a meta-analysis. Review Manager (RevMan)^21^ statistical software, provided by Cochrane, was used to analyze data. Using the random-effects model of meta-analysis we assessed the magnitude of the intervention effects according to study outcomes in two graphs: 1) Hb index of all selected studies, subdivided into food and iron pot / iron ingot subgroups, further stratified to present data on children only using the same subdivisions; 2) prevalence of IDA, subdivided and stratified in the same manner. Data were summarized in a forest-plot type graph.

## Results

### Selection and characterization of studies

A total of 479 studies were retrieved from the databases (226 SciELO/LILACS; 207 MEDLINE (via PubMed); 29 in Cochrane Library; and 17 in EMBASE). Of these 4 duplicate records were removed. All titles and abstracts were reviewed and then 23 articles were read in full and reviewed for eligibility checking. Three articles met inclusion criteria. We also conducted a manual search for citations from the included articles to identify additional relevant studies, a further 8 records were identified. After these reports were screened for eligibility, all studies met inclusion criteria. Thus, eleven RCTs were included in this review (Figure 2).

**Figure 2.**
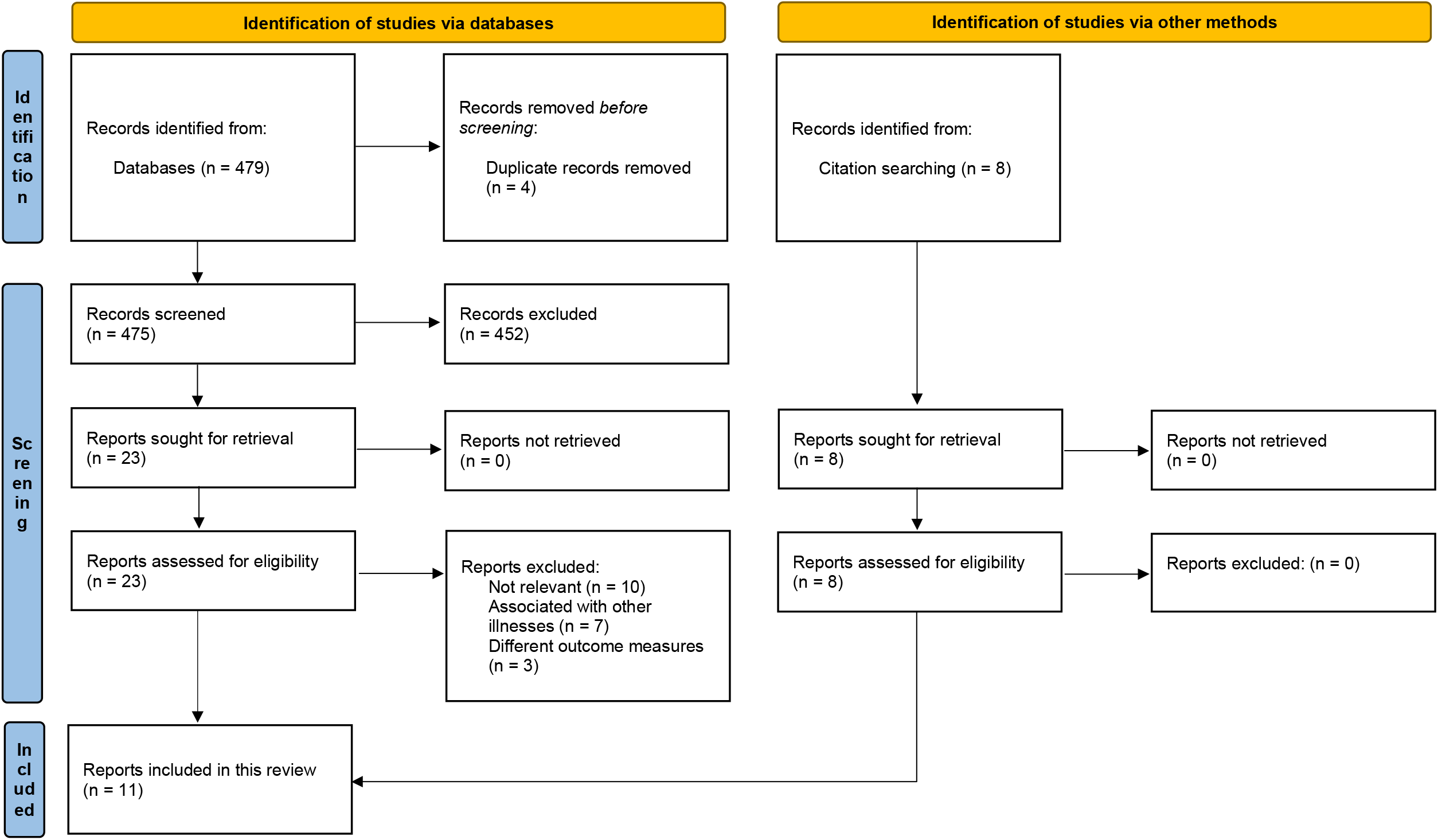
Search strategy according to the PRISMA protocol.^22^

**Figure 3.**
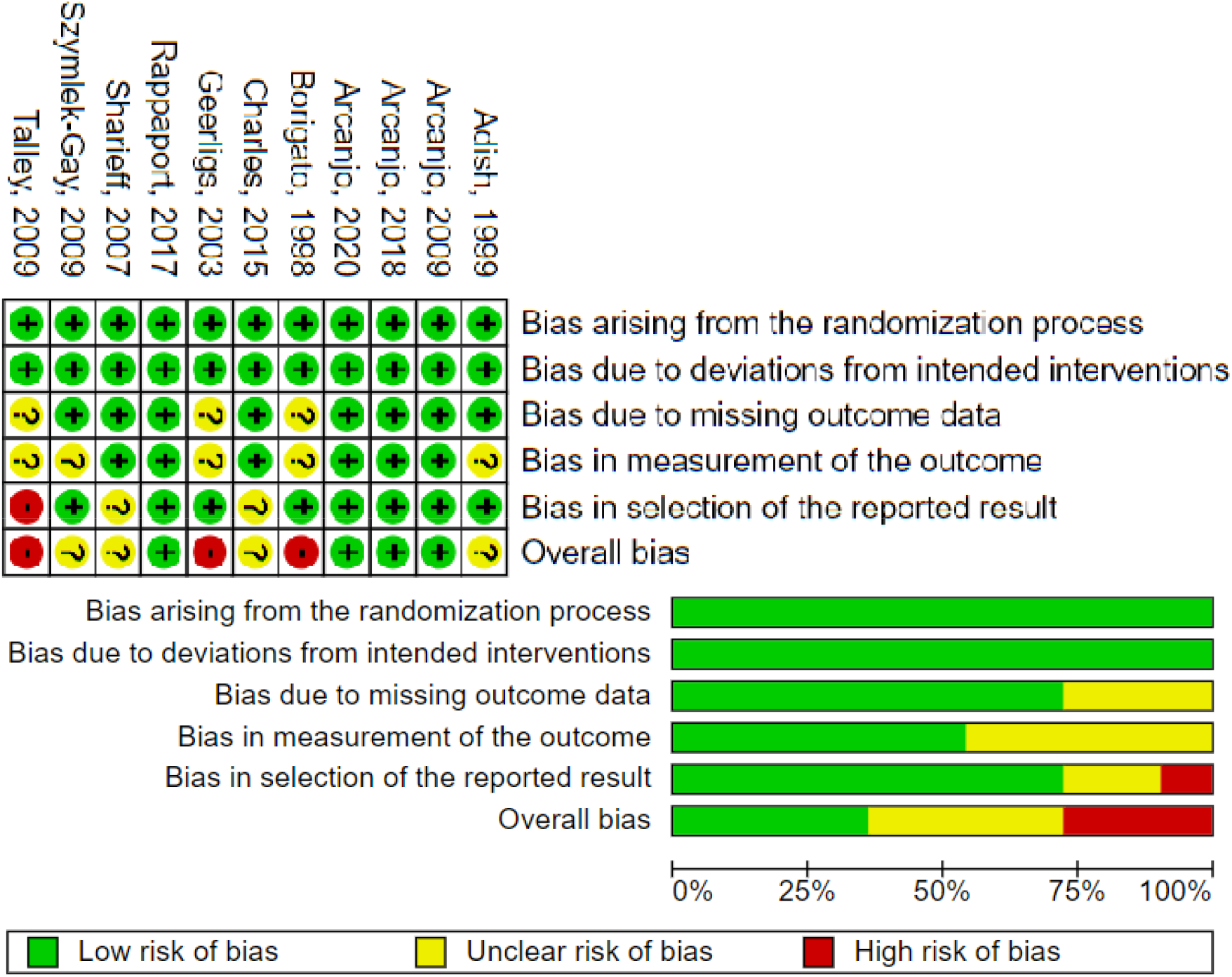
Risk of bias in studies using the Cochrane Risk of Bias tool (RoB 2.0).

### Risk of bias within studies

The quality of the included studies was independently assessed by two researchers (FMBG and FPNA) using the RoB 2.0 tool.^34^ After initial analysis, discrepancies between items were discussed, and consensus was reached. In assessing the risk of bias, all outcomes (primary and secondary) were evaluated. Of the eleven studies evaluated in this review, 4 had low risk of bias, 4 had unclear risk of bias, and 3 had high risk of bias.

### Assessment of the quality of evidence according to GRADE

Included randomized studies had a low risk of bias and two outcomes were directly assessed: 1) differences in Hb levels (11 studies), 2) prevalence of anemia (five studies). Information bias was also unlikely. Regarding the analysis of the two outcomes, there was inconsistency in the results due to the participants’ non-adherence to the proposed interventions. In the iron pots groups, use was not daily (average use was three times a week), and in the red meat group, mean consumption was less than half of what the intervention intended (0.7 dishes/day out of 2 dishes/day). Therefore, we consider the overall certainty of evidence to be moderate (Table 2).

**Table 2.**
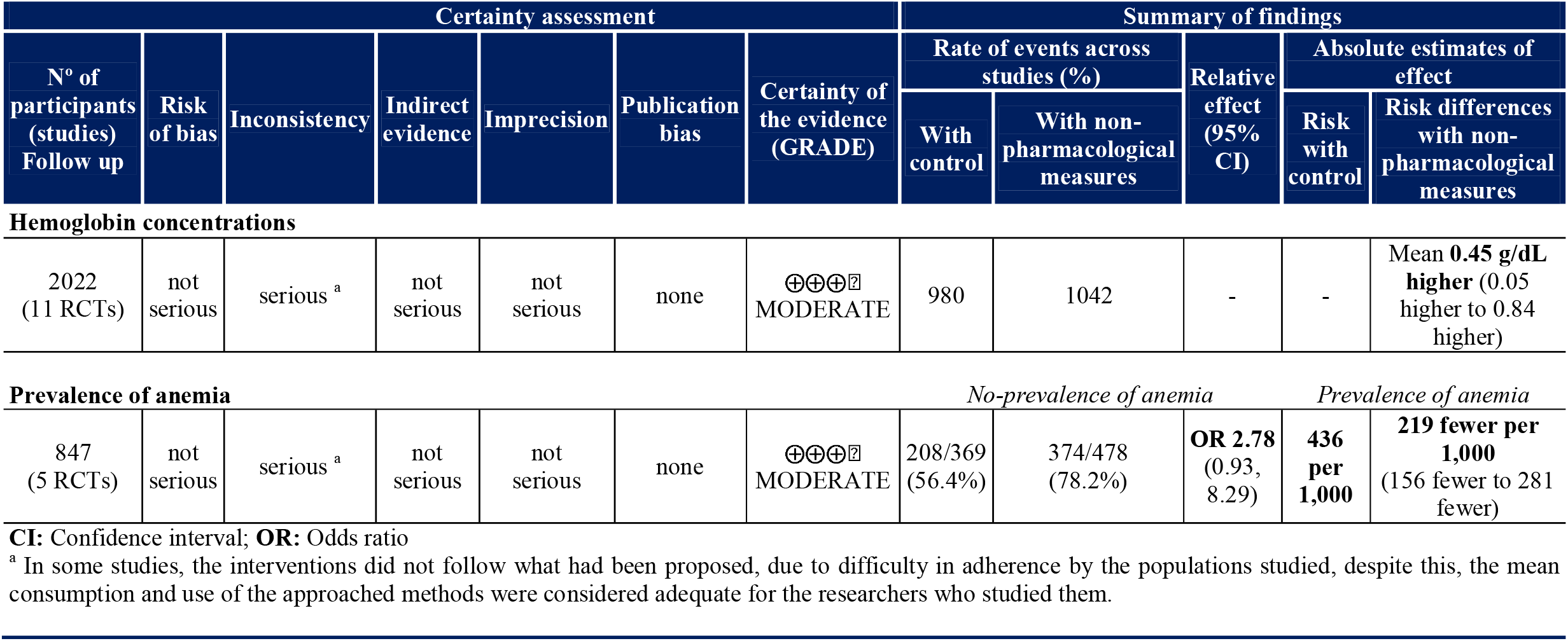
A summary of findings table for evaluating the quality of evidence-non-pharmacological measures compared to control for hemoglobin concentrations and prevalence of anemia according to GRADE.

### Synthesis of results

#### Effect of using food and iron pots / ingots on hemoglobin levels

Estimates showed that the use of non-pharmacological measures was associated with a statistically significant overall increase in mean Hb (mean difference (MD) +0.45 g/dL, 95% CI 0.05 to 0.85, p=0.03), with a high level of heterogeneity among studies (p<0.0001, I^2^=91%). This mean increase in Hb concentrations was also present in the subgroups but without statistical significance (for the studies with food: MD +0.51 g/dL, 95% CI -0.32, 1.34, p=0.22) (for the studies with iron pots / ingots: MD +0.43 g/dL, 95% CI -0.06, 0.92, p=0.09) (Figure 4).

**Figure 4.**
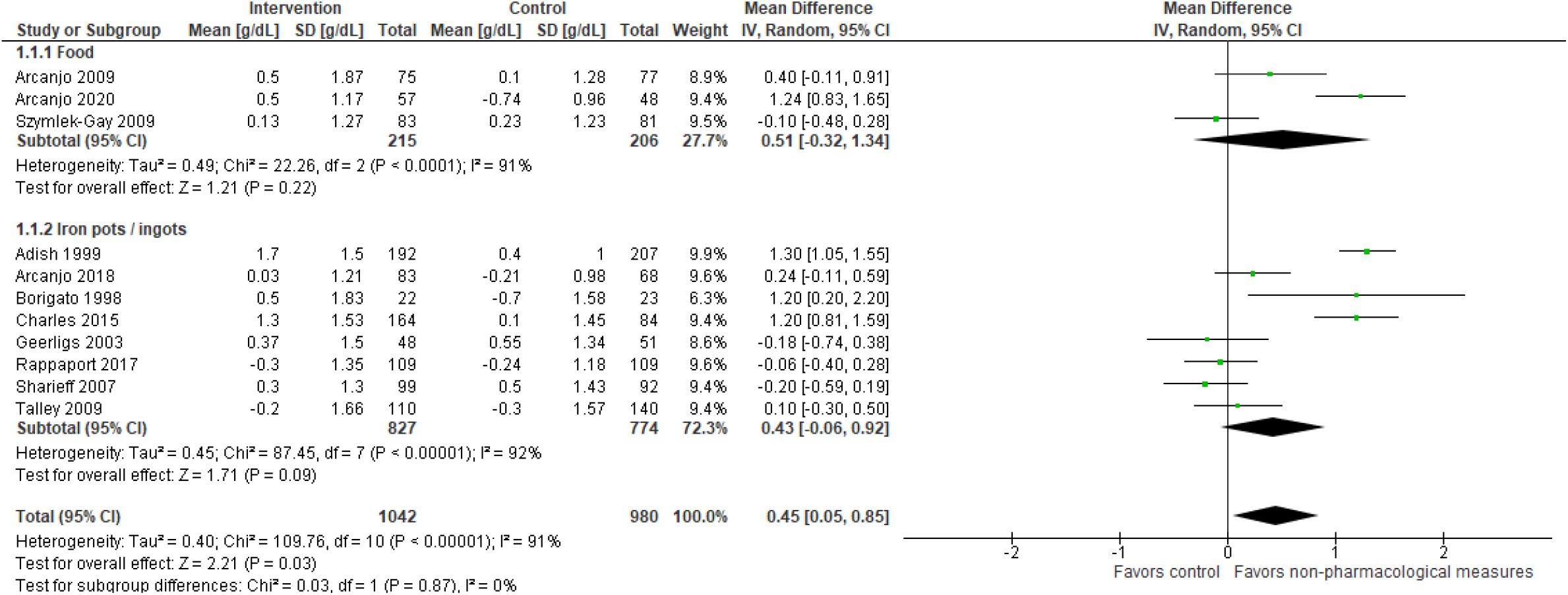
Effect for the overall and subgroup analysis of therapy with non-pharmacological measures on Hb levels.

Considering the studies conducted exclusively with children, the overall effect of non-pharmacological measures on Hb levels was statistically significant (p=0.01), resulting in a mean increase of 0.69 g/dL (95% CI 0.15, 1.24), with a high level of heterogeneity (91%). In the subgroup analysis, only iron pots / ingots produced a significant rise (0.88 g/dL 95% CI 0.05, 1.72, p=0.04) in mean Hb values when compared to food (0.51 g/dL, 95% CI -0.32, 1.34, p=0.22) (Figure 5).

**Figure 5.**
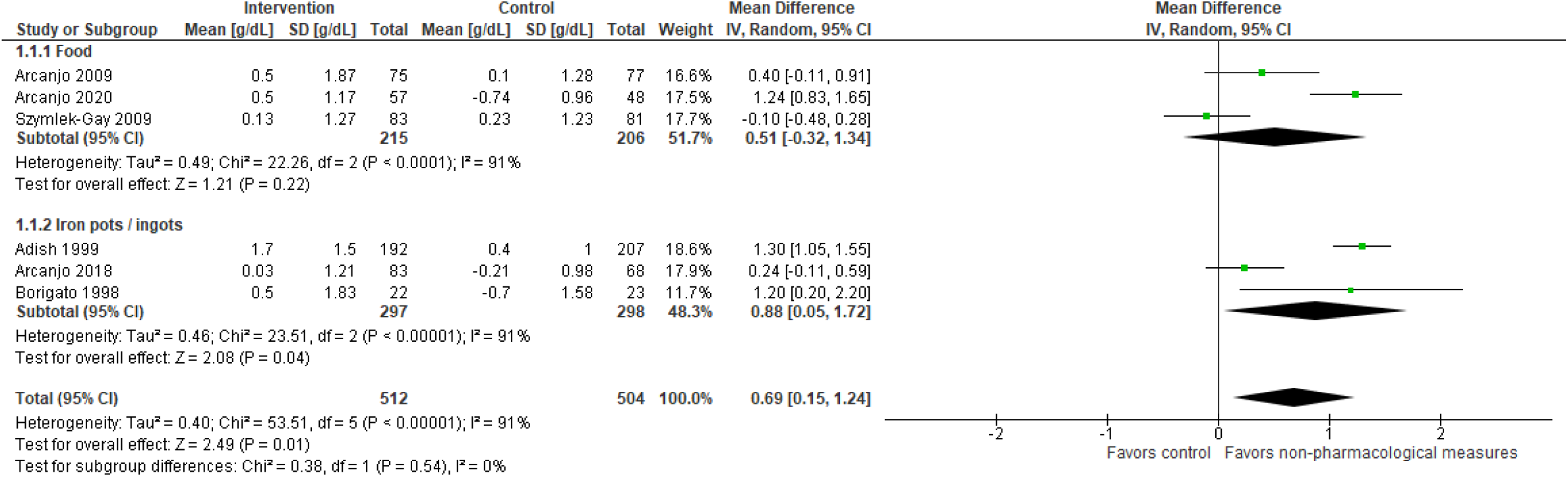
Effect for the overall and subgroup analysis of therapy with non-pharmacological measures on Hb levels in children.

#### Effect of use of food and iron pots / ingots on the prevalence of IDA

The effect of non-pharmacological measures on the prevalence of IDA was analyzed in only 5 RCTs. Participants in the intervention groups were 2.78 times less likely to suffer from IDA than those in the control groups, odds ratio (OR)=2.78, 95% CI 0.93, 8.29, with a high level of heterogeneity (p<0.0001, I^2^=90%), however without significance for the overall effect (p=0.07). In the subgroup analysis, participants in both the food and iron pots / ingots groups were less likely to be anemic when compared to control, OR=4.61, 95% CI 0.75, 28.50 and OR=1.99, 95% CI 0.47, 8.44, respectively, although once again without statistical significance (Figure 6).

**Figure 6.**
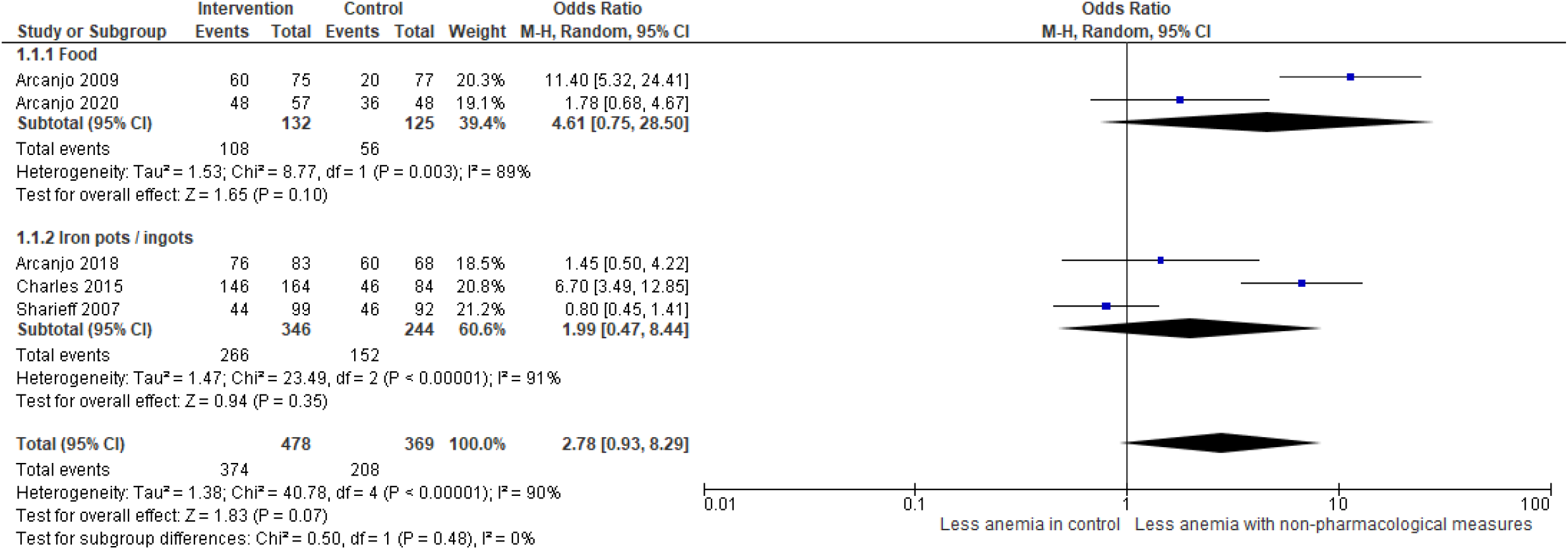
Effect for the overall and subgroup analysis of therapy with non-pharmacological measures on the prevalence of anemia.

When analyzing changes in the prevalence of anemia only in children (3 trials), we found that participants in the interventions group were 3.20 times less likely to have IDA (OR=3.20, 95% CI 0.80, 12.80), without significance (p=0.10). In the food subgroup, IDA was less prevalent in the intervention group, as is the case with iron pots / ingots, although without statistical significance (p=0.10 and 0.50, respectively) (Figure 7).

**Figure 7.**
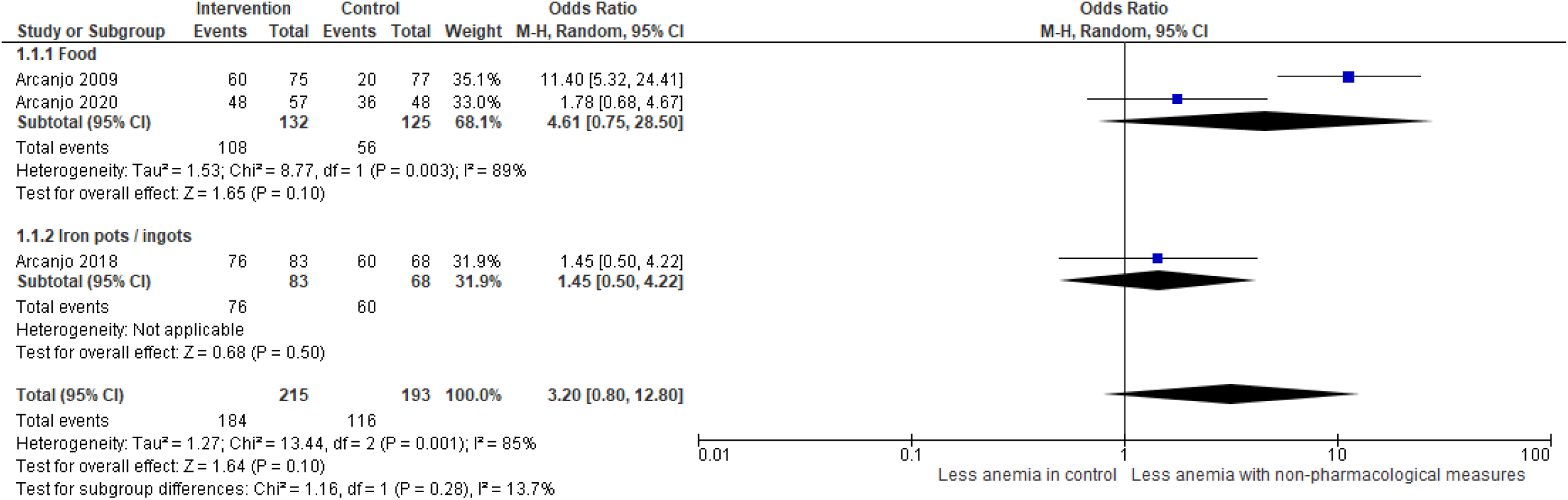
Effect for the overall and subgroup analysis of therapy with non-pharmacological measures on the prevalence of anemia in children.

## Discussion

Our systematic review showed that non-pharmacological treatments for IDA had a significant positive impact on Hb concentrations (MD +0.45 g/dL); however, in the subgroup (food or iron pots / ingots) analysis, although mean Hb values increased this was not statistically significant, probably due to a small subgroup size and a large confidence interval. This interval may result from the heterogeneity of the meta-analysis, by combining studies with different methods, treatments used and target populations, but with a common objective. In the RCTs conducted exclusively with children, again the overall effect of non-pharmacological interventions provided a significant mean increase in Hb concentrations (0.69 g/dL). In the subgroup analyses, we found that iron pots / ingots impacted more significantly on mean Hb values than food (0.88 g/dL vs. 0.51 g/dL) in this population.

For the effect of food interventions on Hb concentrations, only 2 out of 3 interventions provided a positive effect.^28,33^ In the study by Szymlek-Gay et al. (2009),^27^ the authors reported that mean consumption (red meat intervention) was less than half of what the intervention intended (0.7 dishes/day out of 2 dishes/day), they recognized that identifying and removing factors that might have affected adherence could have delivered more effective results. Consequently, this study had a negative impact on the overall effect (MD -0.10, 95% CI -0.48, 0.28) of the studies.

The results of the interventions that used iron pots / ingots for cooking on Hb concentrations were extremely varied, in 5 out of 8 RCTS,^23,24,29,30,32^ the effect of the intervention was superior to that of the control. Although, when analyzing the study by Talley et al. (2009), ^29^ we found that this study was carried out in different aid-dependent refugee camps where the prevalence of anemia had reduced significantly due to the distribution of a corn-soy blend prior to the intervention; furthermore, the authors believe that the response may also be due to lack of compliance with the Fe-alloy pots. The three remaining RCTs^25,26,31^ reported negative effects on Hb values from the use of iron pots / ingots when compared with control. The study by Geerligs et al. (2003),^25^ to assess the effect of cooking in iron pots with aluminum ones, was conducted in rural Malawian households. The authors attributed the inconsistent results of this study to high malaria parasite prevalence in the area. Nevertheless, they maintained the recommendation for this type of intervention. In the two other studies,^26,31^ the results were inconclusive, and the authors recommended further studies on the bioavailability of this iron or the nonuse of these strategies.

However, in studies with children only, the use of iron pots / ingots provided a significant impact on Hb concentrations. For the three studies conducted in children only,^23,24,32^ the authors considered this kind of intervention as a useful adjunct for programs to prevent IDA in at-risk populations, recommending their provision for households in less developed countries.

For the prevalence of anemia, we found that although participants in the intervention groups were less likely to suffer from IDA this probability was weakened due to the small number of trials with this outcome measure, and as consequence we were unable to confirm statistical significance (p=0.07 for all studies, and p=0.10 for studies performed with children). However, this outcome measure was only report in five studies.^26,28,30,32,33^ The results of the food interventions^28,33^ provided a greater impact on the prevalence of anemia, and participants in the intervention groups of these studies were 4.61 times less likely to suffer from IDA. The iron pots / ingots interventions^26,30,32^ presented inclusive findings; in the study by Charles et al. (2015),^30^ participants in the control group were about 4 times more likely to be anemic than those in the intervention groups; for Arcanjo et al. (2018),^32^ there was little difference between the intervention and control groups, (reduction of prevalence of IDA from 10.7% to 8.4% in the intervention group, and 13.2% to 11.8% in the control group); but for Sharieff et al. (2007)^26^ anemia was more prevalent in the intervention group when compared to the control group.

In studies with children only (three RCTs)^28,32,33^ there was a reduction in the prevalence of IDA for studies with food interventions^28,33^; in one^28^ the prevalence of anemia fell from 40% to 20% in the intervention group, with no change in the control group, while in the other^33^ the prevalence of anemia fell from 36.8% to 15.8% in the intervention group, and rose from 16.7% to 25% in the control group. For the study with iron pots,^32^ there was little difference between the intervention and control groups.

### Strengths and limitations of the study

This systematic review evaluated the use of non-pharmacological measures on the prevention and treatment of anemia. These analyses examined the effect of sugar cane honey, brown sugar, red meat and cooking with iron pot or ingots on hemoglobin levels and the prevalence of anemia. To our knowledge, this is the first systematic review and meta-analysis aiming to understand the effect of alternative strategies for the treatment and prevention of IDA.

However, our review has limitations. First, few RCTs have investigated non-pharmacological measures, and the data available for the analysis of the study outcomes was limited. Second, some of the studies were conducted in populations with traditionally low Hb concentrations and high levels of anemia that were possible involved in other nutrition programs at the time or shortly before the interventions. In addition, many of the studies presented large confidence intervals which prevented the authors from making a meaningful interpretation.

## Conclusion

The evidence presented from the eleven RCTs included in this review suggest that these non-pharmacological therapies have a positive effect on iron balance, and can be defined as a useful adjunct to programs to prevent and treat ID in at-risk populations, especially those in low-income environments. The findings from this review show that there are therapeutic alternatives that do not include the use of drugs for the prevention and treatment of diseases related to nutritional deficiencies. The results of this review provide an evidence base for public health managers in the elaboration of public policies focused on this theme.

## Data Availability

All data produced in the present work are contained in the manuscript.

## Abbreviations used in manuscript

GRADE: Grading of Recommendations Assessment, Development and Evaluation
Hb: Hemoglobin
ID: Iron deficiency
IDA: Iron-deficiency anemia
MD: Mean difference
OR: Odds ratio
RCT: Randomized clinical trial
WHO: World Health Organization

## References

1. Soares Magalhães RJ, Clements AC. Spatial heterogeneity of haemoglobin concentration in preschool-age children in sub-Saharan Africa. Bull World Health Organ. 2011;89(6):459–468. doi:10.2471/BLT.10.083568

2. Central Statistical Agency [Ethiopia] and ICF International. Ethiopia Demographic and Health Survey 2011. Addis Ababa, Ethiopia and Calverton, Maryland, USA: Central Statistical Agency and ICF International; 2012.

3. Kassebaum NJ, Jasrasaria R, Naghavi M, Wulf SK, Johns N, Lozano R, et al. A systematic analysis of global anemia burden from 1990 to 2010. Blood. 2014;123(5):615–624. doi:10.1182/blood-2013-06-508325

4. Amarante M, Otigossa A, Sueiro A, Oliveira C, Carvalho S. Anemia Ferropriva: uma visão atualizada. 2015;17(1):34–45.

5. Mujica-Coopman MF, Brito A, López de Romaña D, Ríos-Castillo I, Coris H, Olivares M. Prevalence of Anemia in Latin America and the Caribbean. Food Nutr Bull. 2015;36(2 Suppl):S119–S128. doi:10.1177/0379572115585775

6. Silla LM, Zelmanowicz A, Mito I, Michalowski M, Hellwing T, Shilling MA, et al. High prevalence of anemia in children and adult women in an urban population in southern Brazil. PLoS One. 2013;8(7):e68805. doi:10.1371/journal.pone.0068805

7. Corona LP, Duarte YA, Lebrão ML. Prevalence of anemia and associated factors in older adults: evidence from the SABE Study. Rev Saude Publica. 2014;48(5):723–431. doi:10.1590/s0034-8910.2014048005039

8. Borges RB, Weffort VRS. Anemia no Brasil – revisão [Anemia in Brazil – review]. Rev Med Minas Gerais. 2011;21(3 Supl 1):78–82.

9. Souza KJ, Tabox V de F, Oliveira JMC de, Pierezan MR, Giuffrida R, Bressa RC, et al. Perfil epidemiológico da anemia ferropriva no serviço de hematologia de um hospital público, estado de São Paulo, Brasil. Colloq Vitae. 2013;5(1):18–28. doi: 10.5747/cv.2013.v005.n1

10. Fabian C, Olinto MT, Dias-da-Costa JS, Bairros F, Nácul LC. Prevalência de anemia e fatores associados em mulheres adultas residentes em São Leopoldo, Rio Grande do Sul, Brasil [Anemia prevalence and associated factors among adult women in São Leopoldo, Rio Grande do Sul, Brazil]. Cad Saude Publica. 2007;23(5):1199–1205. doi:10.1590/s0102-311x2007000500021

11. Cançado RD, Chiattone CS. Anemia ferropênica no adulto: causas, diagnóstico e tratamento [Iron deficiency anaemia in the adult: causes, diagnosis and treatment]. Rev Bras Hematol Hemoter. 2010;32(3):240–6. doi: 10.1590/S1516-84842010005000075

12. Lopez A, Cacoub P, Macdougall IC, Peyrin-Biroulet L. Iron deficiency anaemia. Lancet. 2016;387(10021):907–916. doi:10.1016/S0140-6736(15)60865-0

13. Bezerra AGN, Leal VS, Lira PIC de, Oliveira JS, Costa EC, Menezes RCE de, et al. Anemia and associated factors in women at reproductive age in a Brazilian Northeastern municipality. Anemia e fatores associados em mulheres de idade reprodutiva de um município do Nordeste brasileiro. Rev Bras Epidemiol. 2018;21:e180001. doi:10.1590/1980-549720180001

14. World Health Organization. Haemoglobin concentrations for the diagnosis of anaemia and assessment of severity: Vitamin and Mineral Nutrition Information System. Geneva: WHO; 2011. Available from: http://www.who.int/vmnis/indicators/haemoglobin.pdf

15. Silva JBCB da, Vieira GM. Epidemiological profile of anemia in hematology outpatient clinic in private health sector. Rev Med (São Paulo). 2021;100(1):20–7. doi: 10.11606/issn.1679-9836.v100i1p20-27

16. Khoshnevisan F, Kimiagar M, Kalantaree N, Valaee N, Shaheedee N. Effect of nutrition education and diet modification in iron depleted preschool children in nurseries in Tehran: a pilot study. Int J Vitam Nutr Res. 2004;74(4):264–268. doi:10.1024/0300-9831.74.4.264

17. Moher D, Liberati A, Tetzlaff J, Altman DG; PRISMA Group. Preferred reporting items for systematic reviews and meta-analyses: the PRISMA statement. PLoS Med. 2009;6(7):e1000097. doi:10.1371/journal.pmed.1000097

18. Robinson KA, Dickersin K. Development of a highly sensitive search strategy for the retrieval of reports of controlled trials using PubMed. Int J Epidemiol. 2002;31(1):150–153. doi:10.1093/ije/31.1.150

19. Schünemann HJ, Higgins JPT, Vist GE, Glasziou P, Akl EA, Skoetz N, et al. Chapter 14: Completing ‘Summary of findings’ tables and grading the certainty of the evidence. In: Higgins JPT, Thomas J, Chandler J, Cumpston M, Li T, Page MJ, et al. (editors). Cochrane Handbook for Systematic Reviews of Interventions version 6.2 (updated February 2021). London: Cochrane; 2021. Available from www.training.cochrane.org/handbook

20. Guyatt GH, Oxman AD, Vist GE, Kunz R, Falck-Ytter Y, Alonso-Coello P, et al. GRADE: an emerging consensus on rating quality of evidence and strength of recommendations. BMJ. 2008;336(7650):924–926. doi:10.1136/bmj.39489.470347.AD

21. Review Manager (RevMan) [Computer program]. Version 5.3. Copenhagen: The Nordic Cochrane Centre, The Cochrane Collaboration; 2014.

22. Page MJ, McKenzie JE, Bossuyt PM, Boutron I, Hoffmann TC, Mulrow CD, et al. The PRISMA 2020 statement: an updated guideline for reporting systematic reviews. BMJ. 2021;372:n71. doi:10.1136/bmj.n71

23. Borigato EV, Martinez FE. Iron nutritional status is improved in Brazilian preterm infants fed food cooked in iron pots. J Nutr. 1998;128(5):855–859. doi:10.1093/jn/128.5.855

24. Adish AA, Esrey SA, Gyorkos TW, Jean-Baptiste J, Rojhani A. Effect of consumption of food cooked in iron pots on iron status and growth of young children: a randomised trial. Lancet. 1999;353(9154):712–716. doi:10.1016/S0140-6736(98)04450-X

25. Geerligs PP, Brabin B, Mkumbwa A, Broadhead R, Cuevas LE. The effect on haemoglobin of the use of iron cooking pots in rural Malawian households in an area with high malaria prevalence: a randomized trial. Trop Med Int Health. 2003;8(4):310–315. doi:10.1046/j.1365-3156.2003.01023.x

26. Sharieff W, Dofonsou J, Zlotkin S. Is cooking food in iron pots an appropriate solution for the control of anaemia in developing countries? A randomised clinical trial in Benin. Public Health Nutr. 2008;11(9):971–977. doi:10.1017/S1368980007001139

27. Szymlek-Gay EA, Ferguson EL, Heath AL, Gray AR, Gibson RS. Food-based strategies improve iron status in toddlers: a randomized controlled trial12. Am J Clin Nutr. 2009;90(6):1541–1551. doi:10.3945/ajcn.2009.27588

28. Arcanjo FP, Pinto VP, Arcanjo MR, Amici MR, Amâncio OM. Effect of a beverage fortified with evaporated sugarcane juice on hemoglobin levels in preschool children. Rev Panam Salud Publica. 2009;26(4):350–354. doi:10.1590/s1020-49892009001000010

29. Talley L, Woodruff BA, Seal A, Tripp K, Mselle LS, Abdalla F, et al. Evaluation of the effectiveness of stainless steel cooking pots in reducing iron-deficiency anaemia in food aid-dependent populations [published correction appears in Public Health Nutr. 2010 Jan;13(1):145]. Public Health Nutr. 2010;13(1):107–115. doi:10.1017/S1368980009005254

30. Charles CV, Dewey CE, Daniell WE, Summerlee AJ. Iron-deficiency anaemia in rural Cambodia: community trial of a novel iron supplementation technique. Eur J Public Health. 2011;21(1):43–48. doi:10.1093/eurpub/ckp237

31. Rappaport AI, Whitfield KC, Chapman GE, Yada RY, Kheang KM, Louise J, et al. Randomized controlled trial assessing the efficacy of a reusable fish-shaped iron ingot to increase hemoglobin concentration in anemic, rural Cambodian women. Am J Clin Nutr. 2017;106(2):667–674. doi:10.3945/ajcn.117.152785

32. Arcanjo FPN, Macêdo DRR, Santos PR, Arcanjo CPC. Iron Pots for the Prevention and Treatment of Anemia in Preschoolers. Indian J Pediatr. 2018;85(8):625–631. doi:10.1007/s12098-017-2604-x

33. Arcanjo CC, Arcanjo FPN, Teixeira AA, Ponte YMG, Brito GA, Ferreira TP, Nascimento WMC, Santos PR. Sugar Cane Honey is as Effective as Weekly Iron Supplementation to Prevent and Treat Anemia in Preschoolers. International Journal of Health Sciences. 2020 Dez; 8(4):45–53. doi:10.15640/ijhs.v8n4a6

34. Sterne JAC, Savović J, Page MJ, Elbers RG, Blencowe NS, Boutron I, et al. RoB 2: a revised tool for assessing risk of bias in randomised trials. BMJ. 2019;366:l4898. doi:10.1136/bmj.l4898

